# Medical genetics workforce in Brazil: practitioners, services, and disease distribution

**DOI:** 10.1101/2021.10.14.21265027

**Authors:** Carolina Bonilla, Vinicius Albuquerque Sortica, Lavinia Schuler-Faccini, Alicia Matijasevich, Mário C. Scheffer

## Abstract

**Purpose:** In anticipation of the implementation of personalized medicine (PM) in Brazil we assessed the demographic characteristics of its medical genetics workforce together with the distribution of rare genetic diseases (RGD) and hereditary cancer syndromes (HCS) across municipalities in the country.

**Methods:** We used demographic data from an earlier report on medical specialties, and open databases providing summarized data on the public and private healthcare systems, for the years 2019 and 2020. In the public system we considered RGD live births and hospitalizations, and HCS mortality. In the private system we obtained data on RGD, HCS and genetic counselling appointments.

**Results:** The 332 registered medical geneticists (MGs) were mostly female, attended a public medical school, and were predominantly registered in the Southeast. The distribution of MGs overlapped the country-wise distribution of all types of genetic disease and service examined, indicating that ∼30% of the patient population has access to a MG specialist.

**Conclusion:** The Brazilian MG workforce is concentrated in the richest and most populated areas and while it covers a significant proportion of the population there are vast regions with very limited services. The public health system should address these inequalities for a successful transition to PM.

## Introduction

Personalized or precision medicine (PM) has been increasingly promoted as the next desirable step in the pathway towards a customized approach to human health and well-being but the benefits to developing countries are not yet clear^1^. PM was defined by the United States (US) National Institutes of Health (NIH) as the “approach for disease treatment and prevention that takes into account individual variability in genes, environment, and lifestyle for each person”. Essentially, it is proposed that, by focusing on an individual’s genetic background and non-genetic exposures, PM will provide the right treatment, for the right person, at the right time, thus making medical practice more efficient and cost-effective^2^.

It is undeniable that genetic and genomic testing have become more frequent in the clinic since the sequencing of the human genome was finalized in 2003, driven by rapid technological advances and a decline in the cost of genetic tests. However, the fast pace of expansion of genetic assessments in medicine has not been accompanied by a concomitant growth of the medical genetics workforce^3,4^. Genetics personnel shortages have been reported in Europe, the US, and other high-income countries, scarcity that is likely to worsen if no efforts are made to increase the number of medical geneticists and other genetic healthcare providers^4–6^.

Given that these circumstances have been shown to affect developed nations, we were interested in assessing the state of affairs in developing countries, which are likely to produce proportionally less specialized genetics professionals. Therefore, we examined the situation in Brazil, a highly populated country with a strong genetics tradition in medical education and research. Brazil has a free at point of care universal health system, the Sistema Único de Saúde (SUS), that ensures equal access to health care services for its population. In addition, 28% of the population is also covered via private health insurance^7^, which is regulated by the National Agency of Supplementary Health under the umbrella of the Ministry of Health.

This article presents our findings on the characteristics, professional opportunities for, and distribution of medical geneticists (MGs) in relation to demand for services by patients with rare genetic diseases (RGD) and hereditary cancer syndromes (HCS) across Brazil. RGDs are mostly monogenic conditions, which result from highly penetrant modifications in a single gene. Although each disease exists in very low frequencies (affecting 1.3 in 2000 individuals), they cause major impairments to affected subjects, including death. Conversely, HCS include some multifactorial diseases, which are due to the actions of many genes interacting with the environment, and low penetrance mutations of moderate to high frequencies.

As a conclusion from our analysis, we suggest possible avenues through which to expand the medical genetics workforce and/or services to satisfy present needs and in preparation for the growing demand that implementation of PM may require in Brazil.

## Materials and methods

In this analysis we combined data from 2019 and 2020, firstly because these were the latest reports available, but also because 2020 was an atypical year due to the COVID-19 pandemic, and we felt it would not be representative of the situation we wanted to examine. In some cases, information was only available for 2019. Public and private healthcare system databases were accessed during the first semester of 2021. Websites reported were accessed between July and October 2021.

### 1. Characteristics and geographic distribution of the medical genetics workforce

The demographic study of MGs was part of a larger study of medical doctors in Brazil carried out by Scheffer and colleagues^8^. Scheffer et al. used secondary data obtained from the administrative records of the Regional Medical Councils (CRM), the Federal Council of Medicine and the Brazilian Institute of Geography and Statistics (IBGE) census. In Brazil the specialist degree is granted by medical specialty societies, through the Brazilian Medical Association (AMB), or by the Medical Residency programs accredited by the National Commission of Medical Residency (CNRM).

For the analysis of specialist physicians, data from the registration of degrees in the CRMs, the CNRM, and the societies of medical specialties linked to the AMB, was collected. When the geographical distribution of MGs was analysed, specialists with secondary registrations (physicians with an active registration in more than one CRM) were counted in each state where they were acting^8^.

Results are presented as absolute (counts) and relative (percentage) frequencies. Associations between variables were assessed using cross-tabulation tests.

### 2. Job availability

Information about the residency programs in Medical Genetics was extracted from the website of the Brazilian Society of Medical Genetics and Genomics (SBGM, https://www.sbgm.org.br/). Information about reference centres for rare diseases in Brazil was obtained through the platform Many of Us Are Rare (https://muitossomosraros.com.br/), which acts as a provider of original and relevant content on rare diseases for civil society, health authorities, and the media, and the Ministry of Health website (https://www.gov.br/saude/pt-br/composicao/sgtes/doencas-raras/centros-habilitados-para-tratamento-de-doencas-raras). Data collection on SUS facilities with MG positions was obtained from the National Registry of Health Facilities (CNES).

### 3. Distribution of rare genetic diseases and hereditary cancer syndromes

In 2014, national guidelines for the comprehensive care of individuals with rare diseases (Política Nacional de Atenção Integral as Pessoas com Doenças Raras) were created in Brazil. These guidelines classified rare diseases in 4 axes, the first 3 corresponding to genetic diseases: I-congenital malformations, II-intellectual disability, III-metabolic diseases, and IV-non-genetic rare diseases (http://conitec.gov.br/images/Incorporados/DoencasRaras-EixosI-II-III-FINAL.pdf).

In this study, we extracted data from various healthcare databases on diseases falling within the first 3 axes (from here on, rare genetic diseases [RGD]) (Supplementary Tables 1 and 2), and on hereditary cancer syndromes (HCS) described by the US National Institutes of Health (https://rarediseases.info.nih.gov/diseases/diseases-by-category/28/hereditary-cancer-syndromes) and National Cancer Institute (https://www.cancer.gov/about-cancer/causes-prevention/genetics/overview-pdq) (Supplementary Table 3). Given that ∼75% of rare diseases affect children before the age of 5, the addition of HCS was intended as a way of examining genetic counselling attention needed in adulthood, although several HCS also manifest themselves in childhood, therefore some overlapping between these categories is to be expected.

Information on live births and hospital admissions for RGD, and mortality due to HCS within the public system was obtained from the national health information database (DataSUS) harboured online by the Ministry of Health (https://datasus.saude.gov.br/informacoes-de-saude-tabnet/) (Figure 1).

**Figure 1.**
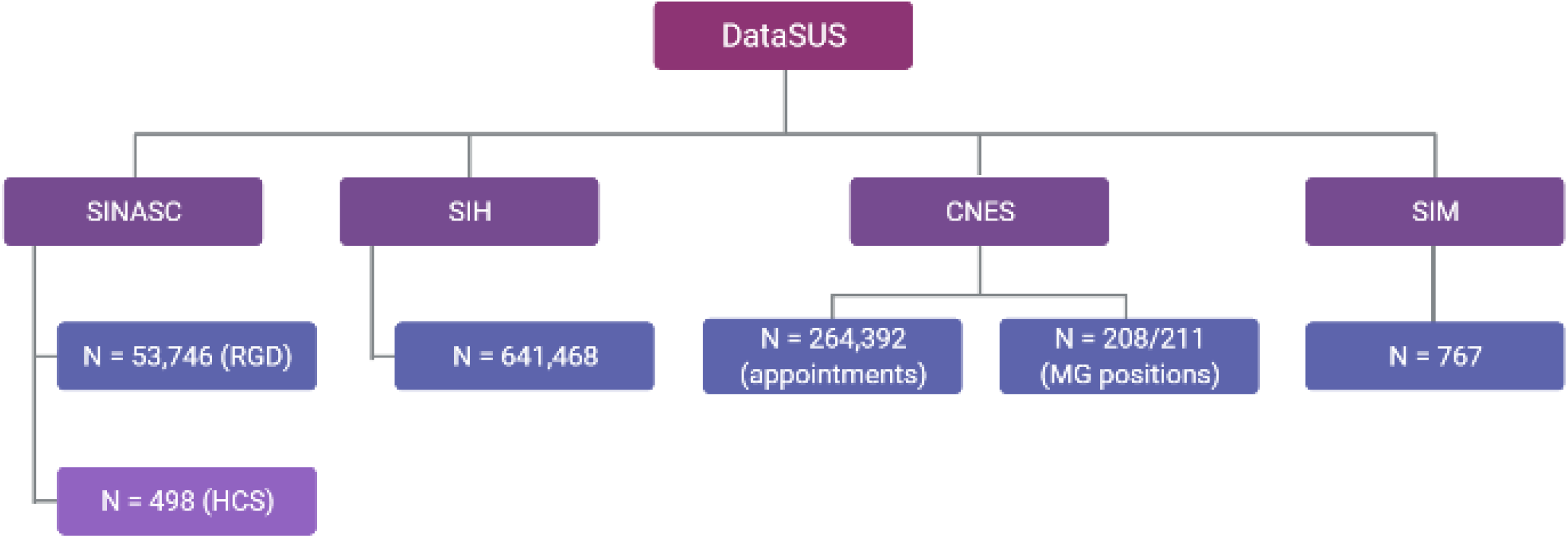
National Health Information database (DataSUS) and subsidiary databases providing information on the public healthcare system used in this study. SINASC = Live Births Information Syste SIH = Hospital Information System CNES = National Registry of Health Facilities SIM = Mortality Information System RGD = rare genetic diseases HCS = hereditary cancer syndromes Figure created with BioRender.com.

Data about live births with RGD and HCS by municipality of residence was extracted from the Live Births Information System (SINASC), which is where congenital abnormalities detectable at birth are officially registered. SINASC relies on the Statement of Live Birth completed at the time of delivery, a mandatory form in Brazil^9^. Some RGD included in the 3 axes listed above were not found in SINASC, those that were, are shown in Supplementary Table 1.

Data on hospitalizations for RGD by municipality of residence was obtained from the Hospital Information System (SIH), while data on care for RGD in the different SUS facilities was obtained from the CNES. To investigate RGD attention at SUS facilities we considered treatment codes 139 (newborn screening service), 145 (clinical laboratory diagnostics) and 168 (care for patients with rare diseases) (Supplementary Table 4). Mortality data for HCS in 2019 by municipality of residence was obtained from the Mortality Information System (SIM) (data for 2020 was unavailable).

In addition, we investigated the number of SUS RGD live births, hospitalizations and HCS deaths, as well as active MGs, in verified clusters of genetic diseases described by Cardoso and colleagues^10^ and the National Census of Isolates (CENISO) at the National Institute of Population Medical Genetics (iNaGeMP, https://www.inagemp.bio.br/), across Brazil, focusing on small municipalities. We also included in this analysis União dos Palmares, a municipality that exhibits high levels of isonymy^11^.

Healthcare data from private providers was acquired from the National Agency for Supplementary Health (ANS) via the Panel for Supplementary Health Information Exchange Standard (D-TISS, https://www.gov.br/ans/pt-br/acesso-a-informacao/perfil-do-setor/dados-e-indicadores-do-setor/d-tiss-painel-dos-dados-do-tiss). This database compiles information on care delivered by health insurance suppliers to their beneficiaries. We extracted information on care for RGD and HCS, and genetic counselling consultations (Supplementary Table 2, Supplementary Figure 1). Only data for 2019 aggregated by state, not by municipality, was available in this system.

Because this study was performed using summarized data from open databases, ethical permissions beyond those secured when the data was originally collected were not required.

## Results

### 1. Characteristics of the medical genetics workforce

In 2019-2020 there were 332 medical geneticists (MGs) in Brazil, 27 more than in 2018^12^, which represents an increase of ∼9%. Sixty-four percent of MGs were female (Table 1). Mean age was 46 years old (range 27-77 years old), while the mean time since graduation was 20.9 years (SD 12.2)^8^. There were no sex differences by age, location or medical school attended (public/private) (Supplementary Table 5).

**Table 1.** Characteristics of the medical genetics workforce in Brazil (2019-2020).

|  | N (332) | % |
| --- | --- | --- |
| **sex** |  |  |
| female | 211 | 63.6 |
| male | 121 | 36.4 |
| **age (years)** |  |  |
| ≤ 29 | 16 | 4.8 |
| 30 – 59 | 255 | 76.8 |
| ≥ 60 | 61 | 18.4 |
| **region where working** |  |  |
| Centre-West | 29 | 8.7 |
| Northeast | 44 | 13.3 |
| North | 5 | 1.5 |
| Southeast | 191 | 57.5 |
| South | 63 | 19.0 |
| **state working location<sup>a</sup>** |  |  |
| capital | 250 | 75.8 |
| interior | 80 | 24.2 |
| **municipality where working (inhabitants)<sup>a</sup>** |  |  |
| > 500,000 | 277 | 83.9 |
| 100,000 – 500,000 | 40 | 12.1 |
| 50,000 – 100,000 | 9 | 2.7 |
| 20,000 – 50,000 | 3 | 0.9 |
| 10,000 – 20,000 | 1 | 0.3 |
| **medical school attended** |  |  |
| private | 100 | 30.1 |
| public | 232 | 69.9 |

<sup>a</sup>Two MGs did not provide municipality location.

Approximately 70% of MGs had graduated from a public medical school (Table 1), although there was a significant difference by age, with older doctors considerably more likely to have graduated from public than private schools (82% of MG ≥ 60 years old vs 50% of MG ≤ 29 years old) (Supplementary Table 5). There was also evidence of a difference by region, i.e. MGs registered in the North and Northeast were more likely to have attended a public medical school than MGs registered in the other regions, particularly the Southeast (Supplementary Table 5).

The schools supplying the highest proportions of MGs were the Federal University of Minas Gerais (5.1%), the Federal University of Rio Grande do Sul (4.8%), the State University of Campinas (4.2%), the University of São Paulo (3.9%), and the Federal University of Paraná (3.9%), all of them public and based in the Southeast and South. There was an imbalance with respect to the location of the medical school where the MGs had graduated and where they ended up working (Supplementary Table 6). For instance, 21% of MGs graduated from a medical school in the state of São Paulo (SP) yet this state hired 34% of the MG workforce. Similarly, the proportion of MGs working in the state of Rio Grande do Sul (RS) doubled the MG output of RS universities. In contrast, the state of Paraná (PR) graduated 17% of MGs but only 5% of them were employed there. We also found that the Southeast and South had more MGs based outside the state capitals compared to the other regions (Supplementary Table 7).

Two-hundred and eight MGs (out of the 252 for whom we had data) had done a residency in Medical Genetics. The second most popular residency program was Paediatrics, which was undertaken by 62 MGs. Eighty-three MGs (25%) had a degree in Paediatrics as well^8^.

### 2. Geographic distribution of the medical genetics workforce

Only 67 of the 5570 municipalities of Brazil had a registered MG in 2019-2020, covering 23 of the 27 federal units (26 states plus the federal district [DF]). The states of Rondônia (RO), Roraima (RR), Amapá (AP) and Tocantins (TO), in the Northern region, do not have MGs working locally. These states as well as four others (Acre [AC], Maranhão [MA], Paraíba [PB] and Rio Grande do Norte [RN]) have no medical schools represented amongst the registered MGs (Supplementary Table 6).

The Southeastern (states of Espírito Santo [ES], Minas Gerais [MG], Rio de Janeiro [RJ] and SP) and Southern (states of PR, Santa Catarina [SC] and RS) regions concentrate the majority of MGs (n = 254, 77%), These regions hold the most populated cities, including São Paulo with ∼12 million people, and constitute the wealthiest regions of the country. As a comparison, the Northern region (AC, Amazonas [AM], AP, Pará [PA], RO, RR and TO), with ∼18 million inhabitants, employs the services of just 5 MGs (1.5%) (Supplementary Table 6).

Additionally, we checked whether recently described clusters of genetic diseases^10^ receive proper MG attention. From the list compiled by the INAGEMP, we assessed clusters that were assigned to a municipality, ignoring those that exist within big cities or across large regions. Despite reporting RGD hospital admissions in the thousands, most population isolates did not have a registered MG (Supplementary Table 8). Thirteen of the 14 available MGs were registered in the municipality of Ribeirão Preto (SP), which, with ∼700,000 inhabitants, is considerably larger than the rest. The municipality of União dos Palmares, representative of a region of high isonymy, also lacked a MG.

### 3. Job availability

#### a. Residency

Brazil has 10 Medical Genetics residency programs, which can be accessed right after undergraduate medical training and open vacancies for ∼23 new trainees each year^13^. The programs last 3 years and are usually offered in the Southeast (n=6), South (n=2), Centre-West (n=1) and Northeast (n=1).

#### b. Other opportunities

MG services at facilities run by SUS were available in 10 different types of facility, out of the 24 that reported providing some kind of genetic service, and out of a total of 43 types of SUS facilities. General hospitals furnished 50% or more of these services, followed by specialized hospitals with over 12%. It should be noted that 208 and 211 MGs of the 332 were accounted for in the SUS facility database in 2019 and 2020, respectively (Supplementary Table 9 and Figure 1).

In addition, 88 reference centres for rare diseases, where it is presumed that MGs are practicing, were described in the Many of Us Are Rare and the Ministry of Health websites. These include public (SUS) and private services and handle non-genetic rare diseases as well. As expected, a majority of reference centres are located in the Southeast (n=49), and the region with the lowest number of them is the Centre-West with 4. The state with the most reference centres is RJ with 20 (Supplementary Table 10).

Considering all information on MGs, SUS facilities with MGs and reference centres together, the number of municipalities served by at least one of these options increases to 111 (2% of all municipalities in the country) (Supplementary Table 11).

### 4. Distribution of rare genetic diseases and hereditary cancer syndromes

#### I. Public healthcare system (DataSUS)

##### Ia. Live births (SINASC)

There were 53,746 live births with an RGD, and 498 live births with an HCS in Brazil in 2019-2020. These represent 1% and 0.01%, respectively, of all live births in the period. 19,033 RGD (35.4%) and 188 HCS (37.8%) occurred in municipalities with an active MG. Their distribution by state and municipality is shown in Table 2 and Supplementary Table 12.

**Table 2.** Distribution of rare genetic diseases (RGD) live births and hospitalizations, and hereditary cancer syndrome (HCS) deaths in the public healthcare system, and medical geneticists (MGs), by state and region (2019-2020).

| region | state | RGD live births 2019-2020 (N = 53,746) | RGD hospital admissions 2019-2020 (N = 641,468) | HCS deaths 2019 (N = 767) | MGs (N = 332) | % population (N=211,755,692)<sup>a</sup> |
| --- | --- | --- | --- | --- | --- | --- |
| North | Rondônia | 427 | 5,596 | 3 | 0 | 0.9 |
| North | Acre | 256 | 1,889 | 3 | 1 | 0.4 |
| North | Amazonas | 850 | 6,259 | 14 | 1 | 2.0 |
| North | Roraima | 199 | 1,885 | 2 | 0 | 0.3 |
| North | Para | 1,718 | 16,244 | 32 | 3 | 4.1 |
| North | Amapá | 583 | 597 | 3 | 0 | 0.4 |
| North | Tocantins | 534 | 3,210 | 2 | 0 | 0.8 |
| total North |  | 4,567 | 35,680 | 59 | 5 | 18,672,591 |
| % North |  | 8.5 | 5.6 | 7.7 | 1.5 | 8.8 |
| Northeast | Maranhão | 1,146 | 16,553 | 35 | 1 | 3.4 |
| Northeast | Piauí | 708 | 8,430 | 9 | 1 | 1.6 |
| Northeast | Ceara | 2,742 | 30,523 | 26 | 9 | 4.3 |
| Northeast | Rio Grande do Norte | 775 | 9,451 | 12 | 1 | 1.7 |
| Northeast | Paraíba | 982 | 8,489 | 9 | 6 | 1.9 |
| Northeast | Pernambuco | 2,741 | 37,012 | 44 | 5 | 4.5 |
| Northeast | Alagoas | 930 | 8,947 | 9 | 5 | 1.6 |
| Northeast | Sergipe | 815 | 4,436 | 12 | 5 | 1.1 |
| Northeast | Bahia | 3,179 | 41,485 | 61 | 11 | 7.1 |
| total Northeast |  | 14,018 | 165,326 | 217 | 44 | 57,374,243 |
| % Northeast |  | 26.1 | 25.8 | 28.3 | 13.3 | 27.2 |
| Southeast | Minas Gerais | 4,239 | 76,984 | 66 | 33 | 10.1 |
| Southeast | Espirito Santo | 1,122 | 12,554 | 18 | 6 | 1.9 |
| Southeast | Rio de Janeiro | 3,084 | 42,799 | 52 | 39 | 8.2 |
| Southeast | São Paulo | 15,944 | 147,297 | 160 | 113 | 21.9 |
| total Southeast |  | 24,389 | 279,634 | 296 | 191 | 89,012,240 |
| % Southeast |  | 45.4 | 43.6 | 38.6 | 57.5 | 42.0 |
| South | Paraná | 2,456 | 49,702 | 63 | 16 | 5.4 |
| South | Santa Catarina | 1,844 | 29,843 | 27 | 6 | 3.4 |
| South | Rio Grande do Sul | 2,701 | 37,578 | 40 | 41 | 5.4 |
| total South |  | 7,001 | 117,123 | 130 | 63 | 30,192,315 |
| % South |  | 13.0 | 18.3 | 17.0 | 19.0 | 14.3 |
| Centre-West | Mato Grosso do Sul | 676 | 6,558 | 7 | 4 | 1.3 |
| Centre-West | Mato Grosso | 702 | 6,661 | 10 | 1 | 1.7 |
| Centre-West | Goiás | 1,661 | 18,591 | 36 | 3 | 3.4 |
| Centre-West | Distrito Federal | 732 | 11,895 | 12 | 21 | 1.4 |
| total Centre-West |  | 3,771 | 43,705 | 65 | 29 | 16,504,303 |
| % Centre-West |  | 7.0 | 6.8 | 8.5 | 8.7 | 7.8 |

<sup>a</sup>Data in this column was obtained from the Brazilian Institute of Geography and Statistics (IBGE) database (https://www.ibge.gov.br/), accessed July 2021.

##### Ib. Hospital admissions (SIH)

In 2019-2020, 641,468 hospital admissions for RGD in the public health system (355,422 in 2019 and 286,046 in 2020) were reported (Figure 1). Of these, 192,407 (30%) took place in the 67 municipalities with a registered MG. The number of hospital admissions for RGD in relation to the number of MGs across states and municipalities is depicted in Table 2, Figure 2 and Supplementary Table 12. We noted that hospitalizations in 2020 were ∼10% lower than in 2019, presumably because of the COVID-19 pandemic.

**Figure 2.**
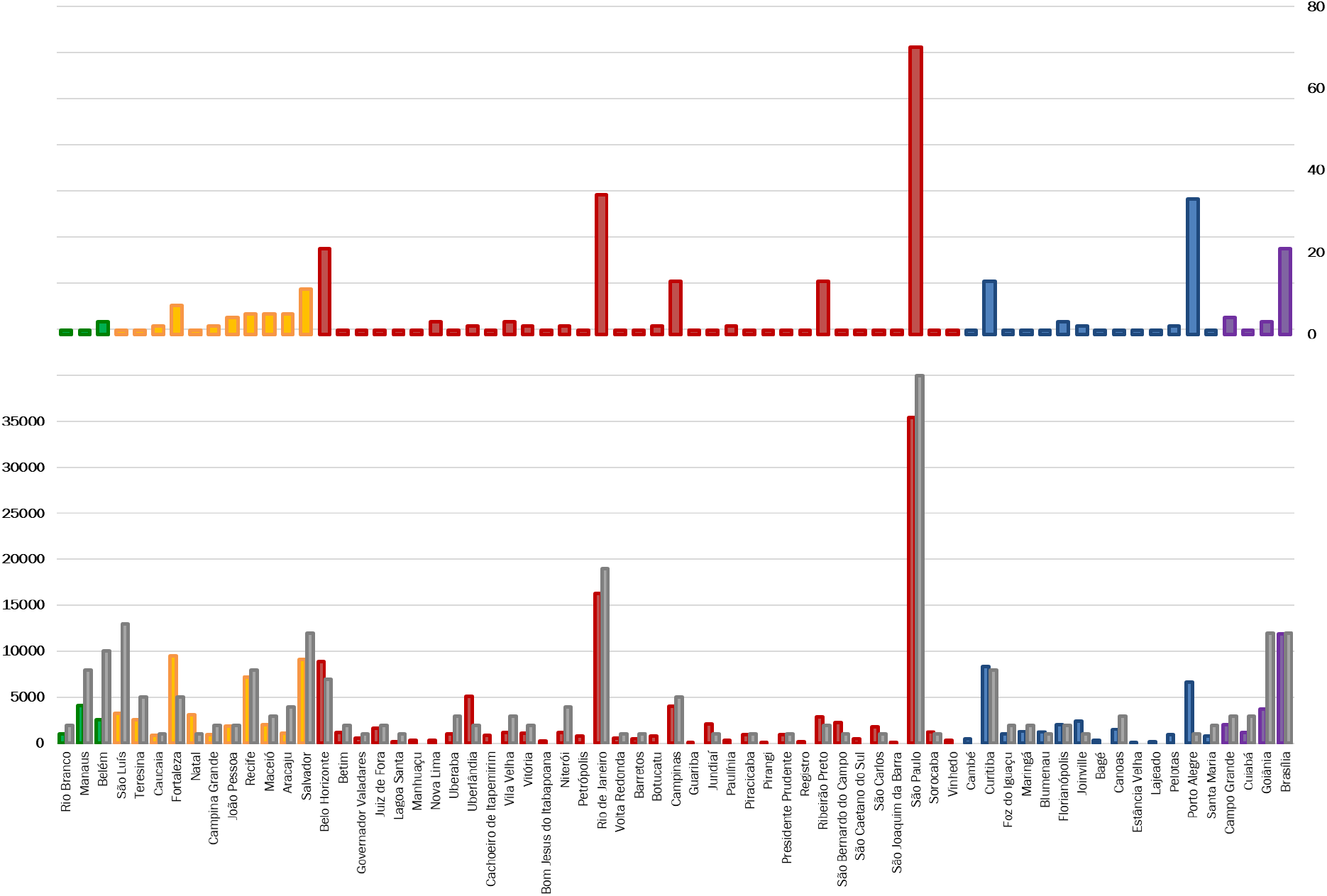
Distribution of medical geneticists, hospital admissions for rare genetic diseases (RGD) and deaths due to hereditary cancer syndromes (HCS) by municipality in Brazil in 2019-2020. Only municipalities with a registered medical geneticist (MG) are shown (n=67). Top panel: registered MGs. Bottom panel: hospital admissions for RGD (coloured) and deaths due to HCS (grey). Green: Northern municipalities. Yellow: Northeastern municipalities. Red: Southeastern municipalities. Blue: Southern municipalities. Purple: Centre-west municipalities. Deaths due to HCS have been multiplied by 1000 for plotting purposes.

Most live births with (45.4%) and hospital admissions for an RGD (43.6%) occurred in the Southeast region, whereas the North exhibited the lowest numbers (8.5% and 5.6%, respectively). The Southeast has proportionally more MGs (57.5%) than RGD live births and hospital admissions, while the opposite is true for the Northeast, which accounted for 26.1% of RGD live births and 25.8% of hospital admissions but is served by 13.3% of MGs, and the North with just 1.5% of MGs (Table 2). At the municipality level, we identified 50 municipalities (3 in the North, 9 in the Northeast, 29 in the Southeast, 8 in the South and 1 in the Centre-West) with over 1000 hospital admissions for RGD in 2019-2020 and no registered MG (Supplementary Table 13).

##### Ic. Mortality (SIM)

Hereditary cancer syndromes were the cause of death of 767 individuals in 2019. In 29.9% of the cases death occurred at a municipality with a registered MG (Supplementary Table 12). The distribution of deaths followed the pattern of live births and hospital admissions mentioned above, namely, the majority taking place in the Southeast (38.6%) and the lowest proportion in the North (7.7%), but the differences between regions were not as large. The number of deaths due to HCS with respect to the number of MGs across municipalities and states is shown in Supplementary Table 12, Figure 2 and Table 2.

We combined the number of RGD hospital admissions and HCS deaths in 2019 to estimate the workload for each MG by municipality (in those municipalities with a registered MG) per year. Patient loads ranged from 21 in the municipality of Pirangi (SP) to 2386 in Manaus (AM), with an average of 496. However, if we calculate the putative load across the country (including all municipalities), that value increases to 1073 patients per MG (i.e. (RGD hospital admissions in 2019 [n = 355,422] + HCS deaths in 2019 [n = 767])/332 MGs). It should be borne in mind that these calculations exclude activities carried out by the same MGs in the private healthcare system.

##### Id. Facilities (CNES)

The analysis of RGD care by SUS facility showed 264,392 appointments reported in 2019-2020 (Figure 1, Supplementary Table 14). The facilities that carried out most of the appointments were the units that provide support for diagnosis and therapy (Unidades de Serviço de Apoio a Diagnose e Terapia [SADT]) with 57.7% of all RGD appointments, followed by general hospitals with 18.6% of these.

##### Ie. National Census of Isolates (CENISO)

The 38 isolates with a confirmed presence of a genetic disease exhibited over 12,000 hospitalizations and 687 live births with a RGD in the time period of interest (Supplementary Table 8). The municipality of União dos Palmares (AL), showing high rates of isonymy, suggestive of endogamic practices^11^, reported 8 RGD live births, 207 RGD hospitalizations, and one HCS death.

#### II. Private healthcare system (D-TISS)

In this system we were able to extract data for RGD-related, genetic counselling, and HCS-related appointments. Numbers were substantially lower than in the public system, with 1,792 RGD-related and 721 HCS-related appointments, although appointments for genetic counselling were much higher (n = 28,171) (Supplementary Figure 1). We could not compare this number to information in DataSUS because the genetic counselling data was incomplete, even though this service is available in the public system. The distribution of these appointments by state with respect to the number of MGs, which was very similar to what was observed in the public system, is shown in Table 3. The vast majority of all types of appointments (65% or more) took place in the Southeast, surpassing the share of active MGs (57.5%). In contrast, the North had proportionally more MGs (1.5%) than its presumed need based on the percentage of appointments (0.1 to 0.6%).

**Table 3.** Distribution of rare genetic disease (RGD), genetic counselling and hereditary cancer syndrome (HCS) consultations in the private healthcare system, and medical geneticists (MGs), by state and region (2019).

| region | state | RGD appointments<br>(N = 1,792) | genetic counselling appointments<br>(N = 28,171) | HCS appointments<br>(N = 721) | MGs (N = 332) |
| --- | --- | --- | --- | --- | --- |
| North | Rondônia | 0 | 0 | 0 | 0 |
| North | Acre | 0 | 0 | 0 | 1 |
| North | Amazonas | 0 | 1 | 1 | 1 |
| North | Roraima | 0 | 0 | 0 | 0 |
| North | Pará | 2 | 19 | 2 | 3 |
| North | Amapá | 0 | 0 | 0 | 0 |
| North | Tocantins | 2 | 0 | 1 | 0 |
| total North |  | 4 | 20 | 4 | 5 |
| % North |  | 0.2 | 0.1 | 0.6 | 1.5 |
| Northeast | Maranhão | 25 | 1 | 4 | 1 |
| Northeast | Piauí | 7 | 0 | 2 | 1 |
| Northeast | Ceará | 6 | 1,796 | 1 | 9 |
| Northeast | Rio Grande do Norte | 4 | 506 | 1 | 1 |
| Northeast | Paraíba | 4 | 11 | 7 | 6 |
| Northeast | Pernambuco | 57 | 1,122 | 17 | 5 |
| Northeast | Alagoas | 3 | 516 | 3 | 5 |
| Northeast | Sergipe | 3 | 7 | 0 | 5 |
| Northeast | Bahia | 58 | 935 | 34 | 11 |
| total Northeast |  | 167 | 4,894 | 69 | 44 |
| % Northeast |  | 9.3 | 17.4 | 9.6 | 13.3 |
| Southeast | Minas Gerais | 126 | 2,692 | 72 | 33 |
| Southeast | Espírito Santo | 33 | 566 | 26 | 6 |
| Southeast | Rio de Janeiro | 378 | 3,392 | 57 | 39 |
| Southeast | São Paulo | 638 | 11,602 | 322 | 113 |
| total Southeast |  | 1175 | 18,252 | 477 | 191 |
| % Southeast |  | 65.6 | 64.8 | 66.2 | 57.5 |
| South | Paraná | 117 | 1,414 | 93 | 16 |
| South | Santa Catarina | 60 | 54 | 17 | 6 |
| South | Rio Grande do Sul | 138 | 1,673 | 36 | 41 |
| total South |  | 315 | 3,141 | 146 | 63 |
| % South |  | 17.6 | 11.2 | 20.3 | 19.0 |
| Centre-West | Mato Grosso do Sul | 18 | 898 | 6 | 4 |
| Centre-West | Mato Grosso | 18 | 184 | 1 | 1 |
| Centre-West | Goiás | 36 | 3 | 6 | 3 |
| Centre-West | Distrito Federal | 59 | 779 | 12 | 21 |
| total Centre-West |  | 131 | 1,864 | 25 | 29 |
| % Centre-West |  | 7.3 | 6.6 | 3.5 | 8.7 |

## Discussion

This study aimed to assess the suitability of the existing medical genetics workforce to satisfy the present needs of the population as well as to anticipate future needs for genetic services if PM is to be implemented in Brazil.

The medical genetics workforce in Brazil is overwhelmingly female, in agreement with the general trend of medical doctors in the country^8^. Remarkably, female MGs are the majority across almost all age categories, including in the group ≥ 60 years old, where a predominance of male doctors is the case for many medical specialties^8^. In a survey of 491 clinical geneticists (MGs actively seeing patients) carried out in the US, 59% of respondents were female, and average age was 51.4 years old, somewhat older than the average of 46.0 years old observed in Brazil^6^.

Participation of private medical schools in MG output is increasing as evidenced by the greater proportion of graduates from private institutions amongst younger MGs, however, 5 public schools still produce almost a quarter of medical doctors that go on to specialize in Medical Genetics. It is also evident that MGs tend to have a strong background in Paediatrics, possibly as a result of the prevalence of RGD in children.

As expected in a country where the richest, most developed and populated regions are in the South and Southeast, concentration of schools with higher numbers of graduates specializing in Medical Genetics, registered MGs, and available workplaces, also occurred in those regions. Equally, a large proportion of medical genetics appointments in the private healthcare system takes place here. However, whilst the distribution of disease outcomes and job offers examined is consistent with the distribution of the population across the country, there is an over-representation of MGs in the Southeast and an under-representation in the North and Northeast, leaving the latter regions underserved, when we consider the public healthcare system (Table 2).

MGs are present in 67 municipalities (1.2% of all municipalities), where ∼28% of the population is based. We determined that overall 30-35% of RGD live births, hospital admissions for RGD, and deaths due to HCS take place in these municipalities. Thus, the deficit of MGs is seemingly not as extensive as it could be. In the same way, several of the municipalities with high numbers of patients and without a registered MG are located in SP state, suggesting that they may be served by MGs who are registered in the big city.

Nevertheless, the absence of MGs in four northern states, with a combined population of almost 5 million people and over 10,000 SUS hospital admissions for RGD in the two-year period, is worrisome. We do not know whether there are MGs carrying out patient consultations remotely or MGs that travel to the area periodically, but even so, addressing the situation in these states should be prioritized.

With respect to job availability, we reported on the number of MGs already present in different SUS facilities, who comprise approximately two thirds of the active MGs. It is interesting to note that the SUS facilities that deliver the largest number of genetics services are the SADTs, yet the general hospitals are the locales where most MGs work.

It is quite clear that the public healthcare system is undertaking a substantial proportion of the tasks of diagnosis, treatment and care of patients with RGD and HCS. However, when it comes to genetic counselling the contribution of private providers appears more significant. Although the available data does not allow us to measure the level of inequality of access of the population to MGs between the public and private subsystems in Brazil, it is known that, in general, people with higher incomes tend to use more specialized medical assistance and those with lower incomes use more generalists and primary care services^14^. Cultural and informational barriers may impact the inequality of access to specialized services, such as those of MGs. In Brazil, access to health services is strongly influenced by the public-private configuration of the health system, social status, income and where people live^15^. Having health insurance reduces financial barriers when consuming specialized services and potentially reduces the waiting time for consultations and exams in medical specialties. The high cost of genetic counselling can make the access be prioritized for those who can afford the expense, either by direct payment (out-of-pocket costs) or through health plans that include these coverages in their contracts. This can detract from the amplification of the offer of MG services in SUS and, with this, hinder collective benefits.

As discussed above, the training of MGs in Brazil is an essentially public investment, financed through taxes and contributions from society as a whole. Most MGs graduated from public medical schools, did their medical residency with public grants and practice in SUS hospitals. However, part of this specialized workforce moves to private services that serve a smaller portion of the population, as attested by the present study with respect to the large number of genetic counselling consultations reported in D-TISS, the private healthcare database.

Our study has strengths and limitations. The status of medical genetics in Brazil has been examined before from a variety of perspectives^13,16–18^, although, as far as we know, this is the first study to include reports of RGD and HCS obtained from open databases in the public and private healthcare systems, in relation to the presence of MGs. The MG demography database was very reliable, since it was created using three other databases that require compulsory registration of medical professionals^8^.

Despite using databases of different origins, which painted a similar global picture, we were limited by the utilisation of secondary data that depends on owner organisations for collection and updating. While we tried to access the most complete and informative outcomes, it is likely that even the data included in this study is affected by underreporting and missing observations. Additionally, other potentially interesting outcomes could not be evaluated because they are not recorded in any of the databases searched (or at all). We should also mention that our description of the contemporary Medical Genetics field in Brazil did not consider the provision of genomic services (for example, whole-genome or whole-exome sequencing), which is growing, especially on private practices, or the genomic proficiency of MGs, elements that will play a key role in the successful implementation of PM.

Based on our findings, we would like to put forward a few suggestions to enhance the reach and performance of MG services in Brazil. These include

a. increasing the number of graduates specialising in Medical Genetics across the country.
b. promoting the specialization in Medical Genetics amongst medical students at universities in the northern states, with the expectation that these professionals will be more likely to settle down in their regions of origin.
c. increasing the number of residency programs in Medical Genetics and the places they offer.
d. encouraging the incursion of MGs into areas of adult-onset Mendelian and multifactorial diseases.
e. adapting job offers to the characteristics of the labour force. For example, since the MG workforce is fundamentally female, positions could be made more flexible and family friendly.
f. supplying healthcare facilities that receive more request for genetic services, such as the SADTs, with additional MG support.
g. implementing MG telemedicine in order to reach more remote locations.
h. broadening genetics education in the undergraduate stages of the medical career, so that general practitioners feel more comfortable treating genetic diseases.
i. expanding the offer of genetics training courses for health professionals in general, not limited to physicians, accessible throughout the country.
j. considering the creation of the genetic counsellor figure, which does not exist in Brazil^13,19^ and does not require a degree in Medicine.

And finally, calling for the careful implementation and systematic updating of public and private healthcare databases so that the picture painted by secondary data analyses is more closely related to reality. Although some of these proposals have been made earlier^13,19–21^, it is important to reconsider them. In addition, mechanisms are also needed to reduce public-private asymmetries within the health system. The added challenge is to bring MGs closer to the SUS and the health needs of most of the population.

In conclusion, we characterized the medical genetics workforce currently active in Brazil, mapped the distribution of genetic diseases as defined by the Ministry of Health, at the municipality level across the country, and uncovered a picture of services in short supply, particularly in the northern regions, that will require serious improvement in the years to come. The country’s progression towards PM, even though the advantages of this route require an appropriate debate that escapes the goals of this study^2,22^, if pursued, should be done by establishing strong foundations that give precedence to the wellbeing of all its inhabitants.

## Data Availability

All data produced in the present study are available upon reasonable request to the authors.

## Acknowledgements

We thank Dr. Alex Cassenote, assistant researcher at the Department of Preventive Medicine, Faculty of Medicine, University of São Paulo, for the development of the medical demography database.

## Funding

CB was supported by a grant from the Pró-Reitoria de Pesquisa of the University of São Paulo (775/2020). AM and MCS receive support from the National Council for Scientific and Technological Development (CNPq), Brazil. VAS is the recipient of a Senior Postdoctoral Fellowship from CNPq (104101/2020-2).

## Supplementary Tables

**Supplementary Table 1**. Rare genetic diseases investigated using databases from the public healthcare system (DataSUS).

**Supplementary Table 2**. Rare genetic diseases investigated using the database for the private healthcare system (D-TISS).

**Supplementary Table 3**. Hereditary cancer syndromes investigated using databases from the public healthcare system (DataSUS).

**Supplementary Table 4**. Attention for rare genetic diseases provided by SUS facilities.

**Supplementary Table 5**. Medical genetics workforce by sex and type of medical school attended.

**Supplementary Table 6**. Distribution of the medical genetics workforce according to medical school attended and current workplace.

**Supplementary Table 7**. Medical genetics workforce by workplace location.

**Supplementary Table 8**. Medical geneticists (MGs), rare genetic diseases (RGD) and hereditary cancer syndromes (HCS) present in clusters of genetic diseases reported by INAGEMP.

**Supplementary Table 9**. Medical geneticists (MGs) working at public healthcare system facilities.

**Supplementary Table 10**. Reference centres for rare diseases listed in the Many of Us are Rare and the Ministry of Health websites.

**Supplementary Table 11**. Municipalities with at least one medical geneticist reported by the medical demography study, one medical geneticist in a SUS facility, or a reference centre for rare diseases.

**Supplementary Table 12**. Distribution of rare genetic diseases (RGD) live births and hospitalizations, and hereditary cancer syndrome (HCS) deaths in the public healthcare system, and medical geneticists (MGs), by municipality (2019-2020).

**Supplementary Table 13**. Municipalities without a registered medical geneticist and a thousand or more hospital admissions for an RGD in 2019-2020.

**Supplementary Table 14**. Attention for rare genetic diseases (RGD) provided by public healthcare system facilities.

## Supplementary Figures

**Supplementary Figure 1**. Database used to extract information from the Brazilian private healthcare system and the outcomes assessed.

RGD = rare genetic diseases

HCS = hereditary cancer syndromes

## Notes

### Competing Interest Statement

Carolina Bonilla is a consultant on ancestry and diversity for the Global Health Equity Advisory Board of Roche/Genentech.

### Funding Statement

CB was supported by a grant from the Pro-Reitoria de Pesquisa of the University of Sao Paulo (775/2020). AM and MCS receive support from the National Council for Scientific and Technological Development (CNPq), Brazil. VAS is the recipient of a Senior Postdoctoral Fellowship from CNPq (104101/2020-2).

### Author Declarations

This study involves only openly available summary human data which can be obtained from DataSUS and D-TISS healthcare databases from Brazil.

## References

1. Mersha TB, Abebe T. Self-reported race/ethnicity in the age of genomic research: its potential impact on understanding health disparities. Hum Genomics. 2015;9(1):1. doi:10.1186/s40246-014-0023-x

2. Iriart JAB. Medicina de precisão/medicina personalizada: análise crítica dos movimentos de transformação da biomedicina no início do século XXI. Cad Saude Publica. 2019;35(3):1–13. doi:10.1590/0102-311×00153118

3. Penon-Portmann M, Chang J, Cheng M, Shieh JT. Genetics workforce: distribution of genetics services and challenges to health care in California. Genet Med. 2020;22(1):227–231. doi:10.1038/s41436-019-0628-5

4. Dragojlovic N, Borle K, Kopac N, et al. The composition and capacity of the clinical genetics workforce in high-income countries: a scoping review. Genet Med. 2020;22(9):1437–1449. doi:10.1038/s41436-020-0825-2

5. Unim B, Pitini E, Lagerberg T, et al. Current Genetic Service Delivery Models for the Provision of Genetic Testing in Europe: A Systematic Review of the Literature. Front Genet. 2019;10(JUN):552. doi:10.3389/fgene.2019.00552

6. Jenkins BD, Fischer CG, Polito CA, et al. The 2019 US medical genetics workforce: a focus on clinical genetics. Genet Med. Published online May 3, 2021. doi:10.1038/s41436-021-01162-5

7. Guilloux AGA, Ramos JA, Citron I, et al. Profiling recent medical graduates planning to pursue surgery, anesthesia and obstetrics in Brazil. BMC Med Educ. 2019;19(1):136. doi:10.1186/s12909-019-1562-6

8. Scheffer M, Cassenote A, Guerra A, et al. Demografía Médica No Brasil 2020.; 2020.

9. Reis LC, Barbian MH, Cardoso-dos-Santos AC, Silva EV de L, Boquett JA, Schuler-Faccini L. Prevalence of congenital anomalies at birth among live births in the state of Maranhão from 2001 to 2016: temporal and spatial analysis. Rev Bras Epidemiol. 2021;24(Suppl 1). doi:10.1590/1980-549720210020.supl.1

10. Cardoso GC, de Oliveira MZ, Paixão-Côrtes VR, Castilla EE, Schuler-Faccini L. Clusters of genetic diseases in Brazil. J Community Genet. 2019;10(1):121–128. doi:10.1007/s12687-018-0369-1

11. Cardoso-dos-Santos AC, Ramallo V, Zagonel-Oliveira M, et al. An invincible memory: what surname analysis tells us about history, health and population medical genetics in the Brazilian Northeast. J Biosoc Sci. 2021;53(2):183–198. doi:10.1017/S0021932020000127

12. Scheffer M. Demografía Médica No Brasil 2018.; 2018.

13. Horovitz DDG, de Faria Ferraz VE, Dain S, Marques-de-Faria AP. Genetic services and testing in Brazil. J Community Genet. 2013;4(3):355–375. doi:10.1007/s12687-012-0096-y

14. Or Z, Jusot F, Yilmaz E. Inégalités de recours aux soins en Europe. Rev économique. 2009;60(2):521. doi:10.3917/reco.602.0521

15. Travassos C, Oliveira EXG de, Viacava F. Desigualdades geográficas e sociais no acesso aos serviços de saúde no Brasil: 1998 e 2003. Cien Saude Colet. 2006;11(4):975–986. doi:10.1590/S1413-81232006000400019

16. Gusmão D, Lessa AC de O, Filho JLT, et al. Perfil clínico-epidemiológico da genética médica no Sistema Único de Saúde: análise do município de São Carlos, SP. Bepa. 2010;7(75):4–15.

17. Iriart JAB, Nucci MF, Muniz TP, Viana GB, Aureliano W de A, Gibbon S. Da busca pelo diagnóstico às incertezas do tratamento: desafios do cuidado para as doenças genéticas raras no Brasil. Cien Saude Colet. 2019;24(10):3637–3650. doi:10.1590/1413-812320182410.01612019

18. Melo DG, Silva AA da, Husny AS El, Ferraz VE de F. Perfil de Competência em Genética para Médicos do Brasil: uma Proposta da Sociedade Brasileira de Genética Médica e Genômica. Rev Bras Educ Med. 2019;43(1 suppl 1):440–450. doi:10.1590/1981-5271v43suplemento1-20180257

19. Ashton-Prolla P, Goldim JR, Vairo FP e., da Silveira Matte U, Sequeiros J. Genomic analysis in the clinic: benefits and challenges for health care professionals and patients in Brazil. J Community Genet. 2015;6(3):275–283. doi:10.1007/s12687-015-0238-0

20. Ferreira CG, Achatz MI, Ashton-Prolla P, Begnami MD, Marchini FK, Stefani SD. Brazilian health-care policy for targeted oncology therapies and companion diagnostic testing. Lancet Oncol. 2016;17(8):e363–e370. doi:10.1016/S1470-2045(16)30171-1

21. Palmero EI, Campacci N, Schüler-Faccini L, et al. Cancer-related worry and risk perception in Brazilian individuals seeking genetic counseling for hereditary breast cancer. Genet Mol Biol. 2020;43(2). doi:10.1590/1678-4685-gmb-2019-0097

22. Novaes HMD, Soárez PC de. Doenças raras, drogas órfãs e as políticas para avaliação e incorporação de tecnologias nos sistemas de saúde. Sociologias. 2019;21(51):332–364. doi:10.1590/15174522-0215121

