## Supplementary Figure 1 for "Medical genetics workforce in Brazil: practitioners, services, and disease distribution"

**Supplementary Figure 1.** Database used to extract information from the Brazilian private healthcare system and the outcomes assessed.

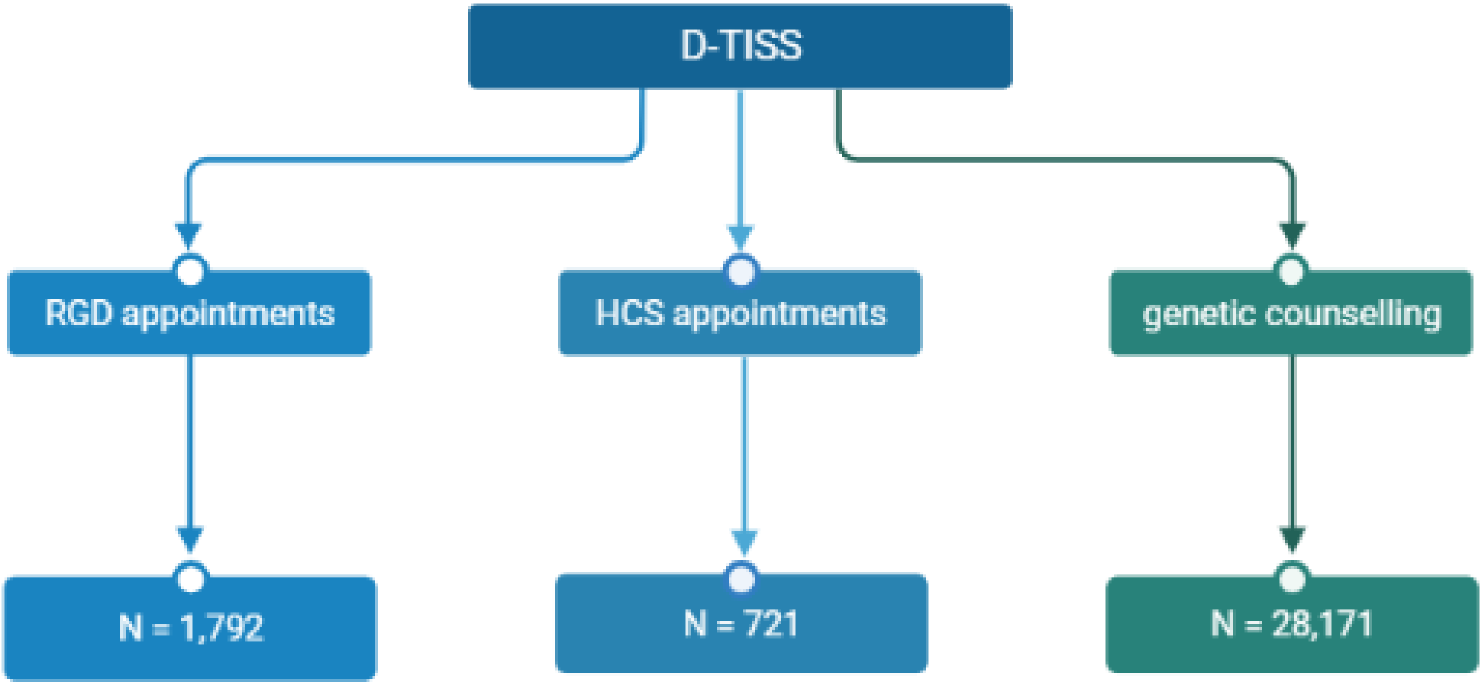


RGD = rare genetic diseases

HCS = hereditary cancer syndromes

Figure created with BioRender.com.
