## Supplementary Tables 1-14 for "Medical genetics workforce in Brazil: practitioners, services, and disease distribution"

**Supplementary Table 1.** Rare genetic diseases investigated using databases from the public healthcare system (DataSUS).

| ICD-10 | disease |
|---|---|
| D80,D80.0,D80.1,D80.2,D80.3,D80.4,D80.5,D80.6D80.7D80.8,D80.9 | immunodeficiency with predominantly antibody defects |
| E51,E52,E53,E54,E55,E56,E58,E59,E60,E61,E63,E64 | other vitamin deficiencies (exclusion: vitamin A deficiency) |
| E70,E71,E72,E73,E74,E75,E76,E77,E78,E79,E80,E83,E84,E85,E86,E87,E88,E89,E90 | metabolic disorders (exclusion: androgen resistance syndrome, congenital adrenal hyperplasia, Ehlers-Danlos syndrome, haemolytic anaemias due to enzyme disorders, Marfan syndrome, 5-alpha-reductase deficiency) |
| F70,F71,F72,F73,F78,F79,F80,F81,F82,F83,F84,F88,F89,F90 | mental retardation |
| F70,F71,F72,F73,F78,F79,F80,F81,F82,F83,F84,F88,F89,F90,F99 | disorders of psychological development and unspecified mental disorder |
| G90,G91,G92,G93,G94,G95,G96,G97,G98,G99 | other disorders of the nervous system |
| Q05 | spina bifida |
| Q00,Q01,Q02,Q03,Q04,Q05,Q06,Q07 | congenital malformations of the nervous system |
| Q20,Q21,Q22,Q23,Q24,Q25,Q26,Q27,Q28 | congenital malformations of the circulatory system |
| Q35,Q36,Q37 | cleft lip and cleft palate |
| Q38,Q39,Q40,Q41,Q42,Q43,Q44,Q45 | other congenital malformations of the digestive system |
| Q50,Q51,Q52,Q53,Q54,Q55,Q56,Q60,Q61,Q62,Q63,Q64 | Congenital malformations of genital organs and urinary system |
| Q65,Q66,Q67,Q68,Q69,Q70,Q71,Q72,Q73,Q74,Q75,Q76,Q77,Q78,Q79 | congenital malformations and deformations of |

| | |
|---|---|
| | the musculoskeletal system |
| Q80,Q81,Q82,Q83,Q84,Q85,Q86,Q87,Q89 | other congenital malformations |
| Q90,Q91,Q92,Q93,Q95,Q96,Q97,Q98,Q99 | chromosomal abnormalities, not elsewhere classified |

In red: RGD and HCS reported by SINASC as well.

**Supplementary Table 2.** Rare genetic diseases investigated using the database for the private healthcare system (D-TISS).

| ICD-10 | disease |
|---|---|
| D82.0 | Wiskott-Aldrich syndrome |
| D82.1 | Di George syndrome |
| E16.1 | other hypoglycaemia |
| E53 | deficiency of other B group vitamins |
| E70 | disorders of aromatic amino-acid metabolism |
| E71 | disorders of branched-chain amino-acid metabolism and fatty-acid metabolism |
| E71.0 | maple-syrup-urine disease |
| E71.1 | other disorders of branched-chain amino-acid metabolism |
| E71.3 | disorders of fatty-acid metabolism |
| E72 | other disorders of amino-acid metabolism |
| E72.0 | disorders of amino-acid transport |
| E72.1 | disorders of sulfur-bearing amino-acid metabolism |
| E72.2 | disorders of urea cycle metabolism |
| E72.3 | disorders of lysine and hydroxylysine metabolism |
| E72.4 | disorders of ornithine metabolism |
| E72.5 | disorders of glycine metabolism |
| E74 | other disorders of carbohydrate metabolism |
| E75 | disorders of sphingolipid metabolism and other lipid storage disorders |
| E75.0 | $GM_2$ gangliosidosis |
| E75.1 | other gangliosidosis |
| E76 | disorders of glycosaminoglycan metabolism |
| E77 | disorders of glycoprotein metabolism |
| E77.0 | defects in post-translational modification of lysosomal enzymes |
| E77.1 | defects in glycoprotein degradation |
| E77.8 | other disorders of glycoprotein metabolism |
| F70 | mild mental retardation |
| F71 | moderate mental retardation |
| F72 | severe mental retardation |

| | |
|---|---|
| F73 | profound mental retardation |
| F78 | other mental retardation |
| F79 | unspecified mental retardation |
| F84.0 | pervasive developmental disorders |
| G12.0 | infantile spinal muscular atrophy, type I [Werdnig-Hoffman] |
| G12.1 | other inherited spinal muscular atrophy |
| G12.2 | motor neuron disease |
| G37 | other demyelinating diseases of central nervous system |
| G71.0 | muscular dystrophy |
| G71.1 | myotonic disorders |
| G71.2 | congenital myopathies |
| G71.3 | mitochondrial myopathy, not elsewhere classified |
| G72 | other myopathies |
| Q00 | anencephaly and similar malformations |
| Q85 | phakomatoses, not elsewhere classified |
| Q85.0 | neurofibromatosis (non-malignant) |
| Q85.1 | tuberous sclerosis |
| Q85.8 | other phakomatoses, not elsewhere classified |
| Q87 | other specified congenital malformation syndromes affecting multiple systems |
| Q87.0 | congenital malformation syndromes predominantly affecting facial appearance |
| Q87.1 | congenital malformation syndromes predominantly associated with short stature |
| Q87.8 | other specified congenital malformation syndromes, not elsewhere classified |
| Q91.0 a Q91.3 | trisomy 18 |
| Q91.4 a Q91.5 | trisomy 13 |
| Q92.2 | major partial trisomy |
| Q92.3 | minor partial trisomy |
| Q92.6 | extra marker chromosomes |
| Q92.8 | other specified trisomies and partial trisomies of autosomes |
| Q92.9 | trisomy and partial trisomy of autosomes, unspecified |
| Q93.3 | deletion of short arm of chromosome 4 |
| Q93.5 | other deletions of part of a chromosome |
| Q93.9 | deletion from autosomes, unspecified |
| Q96 | Turner syndrome |
| Q99 | other chromosome abnormalities, not elsewhere classified |
| Q99.0 | chimera 46,XX/46,XY |
| Q99.2 | fragile X chromosome |
| Q99.8 | other specified chromosome abnormalities |

**Supplementary Table 3.** Hereditary cancer syndromes investigated using databases from the public healthcare system (DataSUS).

| ICD-10 | disease |
|---|---|
| B07 | viral warts |
| C96.0 | multifocal and multisystemic (disseminated) Langerhans-cell histiocytosis [Letterer-Siwe disease] |
| C96.5 | multifocal and unisystemic Langerhans-cell histiocytosis |
| C96.6 | unifocal Langerhans-cell histiocytosis |
| C97 | malignant neoplasms of independent (primary) multiple sites |
| Q43.1 | Hirschsprung disease |
| Q81 | epidermolysis bullosa |
| Q81.1 | epidermolysis bullosa letalis |
| Q81.2 | epidermolysis bullosa dystrophica |
| Q81.8 | other epidermolysis bullosa |
| Q81.9 | epidermolysis bullosa, unspecified |
| Q82.8 | other specified congenital malformations of skin |
| Q82.1 | xeroderma pigmentosum |
| Q85.0 | neurofibromatosis (non-malignant) |
| Q85.1 | tuberous sclerosis |
| Q85.8 | other phakomatoses, not elsewhere classified |
| Q87.0 | congenital malformation syndromes predominantly affecting facial appearance |
| Q87.3 | congenital malformation syndromes involving early overgrowth |
| Q87.8 | other specified congenital malformation syndromes, not elsewhere classified |

**Supplementary Table 4.** Attention for rare genetic diseases provided by SUS facilities.

| code | DataSUS service classification |
|---|---|
| 139 | *newborn screening service* |
| 001 | treatment of newborns with hypothyroidism and phenylketonuria |
| 002 | treatment of newborns with sickle cell disease |
| 003 | treatment of newborns with cystic fibrosis |
| 004 | treatment of newborns with other congenital disorders |
| 005 | monitoring patients diagnosed with congenital adrenal hyperplasia |
| 145 | *clinical laboratory diagnostic service* |
| 011 | genetic testing |
| 012 | tests for newborn screening |
| 168 | *care for patients with rare diseases* |
| 001 | specialized care in rare diseases |
| 002 | reference in rare diseases |

**Supplementary Table 5.** Medical genetics workforce by sex and type of medical school attended.

| | sex | | medical school | | | |
|---|---|---|---|---|---|---|
| | | | private | | public | |
| **sex** | **N (F/M)** | **% F** | **N** | **%** | **N** | **%** |
| female | - | - | 67 | 67.0 | 144 | 62.1 |
| male | - | - | 33 | 33.0 | 88 | 36.5 |
| p-value | | | 0.39 | | | |
| **age (years)** | | | | | | |
| ≤ 29 | 8/8 | 50.0 | 8 | 50.0 | 8 | 50.0 |
| 30 – 59 | 167/88 | 65.5 | 81 | 31.8 | 174 | 68.2 |
| ≥ 60 | 36/25 | 59.0 | 11 | 18.0 | 50 | 82.0 |
| p-value | 0.33 | | 0.02 | | | |
| **region where working** | | | | | | |
| Centre-West | 20/10 | 66.7 | 8 | 26.7 | 22 | 73.3 |
| Northeast&North[a] | 31/15 | 67.4 | 7 | 15.2 | 39 | 84.8 |
| Southeast | 119/75 | 61.3 | 70 | 36.1 | 124 | 63.9 |
| South | 41/21 | 66.1 | 15 | 24.2 | 47 | 75.8 |
| p-value | 0.80 | | 0.03 | | | |

[a]The northeast and northern regions were combined due to the North not having any private school graduates.

F = female, M = male.

**Supplementary Table 6.** Distribution of the medical genetics workforce according to medical school attended and current workplace.

| region/state | medical school | | work | |
|---|---|---|---|---|
| | N | % | N | % |
| **North** | 4 | 1.2 | 5 | 1.5 |
| Rondônia - RO | 0 | 0 | 0 | 0 |
| Acre - AC | 0 | 0 | 1 | 0.3 |
| Amazonas - AM | 2 | 0.6 | 1 | 0.3 |
| Roraima - RR | 0 | 0 | 0 | 0 |
| Para - PA | 2 | 0.6 | 3 | 0.9 |
| Amapá - AP | 0 | 0 | 0 | 0 |
| Tocantins - TO | 0 | 0 | 0 | 0 |
| **Northeast** | 24 | 7.2 | 44 | 13.3 |
| Maranhão - MA | 0 | 0 | 1 | 0.3 |
| Piauí - PI | 2 | 0.6 | 1 | 0.3 |
| Ceará - CE | 2 | 0.6 | 9 | 2.7 |
| Rio Grande do Norte - RN | 0 | 0 | 1 | 0.3 |
| Paraíba - PB | 0 | 0 | 6 | 1.8 |
| Pernambuco - PE | 3 | 0.9 | 5 | 1.5 |
| Alagoas - AL | 1 | 0.3 | 5 | 1.5 |

| | | | | |
|---|---|---|---|---|
| Sergipe - SE | 4 | 1.2 | 5 | 1.5 |
| Bahia - BA | 12 | 3.6 | 11 | 3.3 |
| **Southeast** | 165 | 49.7 | 191 | 57.5 |
| Minas Gerais - MG | 46 | 13.9 | 33 | 9.9 |
| Espirito Santo - ES | 6 | 1.8 | 6 | 1.8 |
| Rio de Janeiro -RJ | 42 | 12.7 | 39 | 11.8 |
| São Paulo - SP | 71 | 21.4 | 113 | 34.0 |
| **South** | 85 | 25.6 | 63 | 19.0 |
| Paraná - PR | 56 | 16.9 | 16 | 4.8 |
| Santa Catarina - SC | 7 | 2.1 | 6 | 1.8 |
| Rio Grande do Sul - RS | 22 | 6.6 | 41 | 12.4 |
| **Centre-West** | 54 | 16.3 | 29 | 8.7 |
| Mato Grosso do Sul - MS | 5 | 1.5 | 4 | 1.2 |
| Mato Grosso - MT | 12 | 3.6 | 1 | 0.3 |
| Goiás - GO | 12 | 3.6 | 3 | 0.9 |
| Distrito Federal - DF | 25 | 7.5 | 21 | 6.3 |

**Supplementary Table 7.** Medical genetics workforce by workplace location.

| | **capital** | | **interior** | |
|---|---|---|---|---|
| **region where working** | **N** | **%** | **N** | **%** |
| Centre-West | 28 | 93.3 | 2 | 6.7 |
| Northeast&North[a] | 43 | 93.5 | 3 | 6.5 |
| Southeast | 131 | 67.9 | 62 | 32.1 |
| South | 48 | 78.7 | 13 | 21.3 |
| p-value | $1.93 \times 10^{-4}$ | | | |

[a]The northeast and northern regions were combined due to the North not having any MGs working in cities outside of state capitals.

**Supplementary Table 8.** Medical geneticists (MGs), rare genetic diseases (RGD) and hereditary cancer syndromes (HCS) present in clusters of genetic diseases reported by iNaGeMP (accessed July 2021).

| state | municipality | genetic disease | MIM code[a] | aetiology | RGD live births 2019-2020 | RGD hospitalizations 2019-2020 | HCS deaths 2019 | MGs |
|---|---|---|---|---|---|---|---|---|
| MA | Cururupu | Oculocutaneous albinism type II | 203200 | AR | 4 | 106 | 0 | 0 |
| CE | Aracati | Multiple familial trichoepithelioma type 1 | 601606 | AD | 29 | 133 | 1 | 0 |
| CE | Crateús | Spinocerebellar ataxia type 7 (SCA 7) | 164500 | AD | 11 | 804 | 0 | 0 |
| CE | Tabuleiro do Norte | Gaucher disease type I | 230800 | AR | 8 | 167 | 0 | 0 |
| RN | Riacho de Santana | Santos syndrome | 613005 | AR | 3 | 0 | 0 | 0 |
| RN | São Miguel | General congenital lipodystrophy type 2 | 269700 | AR | 0 | 48 | 0 | 0 |
| RN | Serrinha dos Pintos | Spastic paraplegia, optic atrophy and neuropathy (SPOAN) | 609541 | AR | 0 | 5 | 0 | 0 |
| PB | Gado Bravo | Usher syndrome type I | 276900 | AR | 1 | 19 | 0 | 0 |
| PB | Lagoa | Consanguinity with increased prevalence of disabilities (mental or physical) | | MF | 1 | 5 | 0 | 0 |
| PE | Fernando de Noronha | Alzheimer's disease | | MF | 3 | 12 | 0 | 0 |
| PE | Gameleira | Short-rib thoracic dysplasia 2 with or without polydactyly | 611263 | AR | 6 | 53 | 0 | 0 |
| | | Verma-Namouff syndrome | 613091 | AR | | | | |
| PE | Orobó | Laron syndrome | 262500 | AR | 3 | 61 | 0 | 0 |
| AL | Água Branca | Aniridia | 106210 | AD | 11 | 37 | 0 | 0 |
| AL | Craíbas | Consanguinity and skeletal disorder, etiology not identified yet | | NI | 6 | 76 | 0 | 0 |
| AL | Feira Grande | Huntington disease | 143100 | AD | 2 | 64 | 0 | 0 |
| AL | Maravilha | Unknown genodermatosis | | AR | 6 | 21 | 0 | 0 |
| AL | Mata Grande | Chondrodysplasia, Blomstrand Type | 215045 | AR | 11 | 8 | 0 | 0 |

| | | | | | | | | |
|---|---|---|---|---|---|---|---|---|
| AL | União dos Palmares | high isonymy | | | 8 | 207 | 1 | 0 |
| SE | Itabaianinha | Isolated growth hormone deficiency type IA | 262400 | AR | 10 | 68 | 0 | 0 |
| BA | Barra da Estiva | Epidermolysis bullosa | | AR | 3 | 88 | 2 | 0 |
| BA | Livramento de Nossa Senhora | Epidermolysis bullosa | | AR | 12 | 101 | 0 | 0 |
| BA | Monte Santo | Deafness | 220290 | AR | 5 | 69 | 0 | 0 |
| | | Mucopolysaccharidosis type VI | 253200 | AR | | | | |
| BA | Vitória da Conquista | Epidermolysis bullosa | | AR | 76 | 1427 | 0 | 0 |
| MG | Alfenas | Oral clefts | | MF | 10 | 867 | 0 | 0 |
| MG | Bueno Brandão | Osteogenesis imperfecta type VI (OI6) | 613982 | AR | 3 | 35 | 0 | 0 |
| MG | Ervália | Huntington's disease | 143100 | AD | 2 | 60 | 0 | 0 |
| MG | Guaxupé | Multiple endocrine neoplasia type 1 (MEN1) | 131100 | AD | 9 | 342 | 0 | 0 |
| MG | Pouso Alegre | Neu-Laxova syndrome 1 (NLS1) | 256520 | AR | 28 | 374 | 0 | 0 |
| RJ | Duque de Caxias | Aggressive periodontitis type 1 | 170650 | AR | 268 | 2196 | 3 | 0 |
| SP | Indaiatuba | Dandy-Walker syndrome (DWS) | 220200 | AR | 35 | 494 | 1 | 0 |
| SP | Jacupiranga | Essential hypertension and consanguinity | 145500 | MF | 1 | 69 | 1 | 0 |
| SP | Mococa | Multiple endocrine neoplasia type 1 (MEN1) | 131100 | AD | 24 | 397 | 0 | 0 |
| SP | Ribeirão Preto | Gomez-Lopez-Hernandez syndrome | 601853 | AR | 296 | 2934 | 2 | 13 |
| SP | | Spinocerebellar ataxia type 1 (SCA1) | 164400 | AD | | | | |
| SP | Vinhedo | Fraser syndrome | 219000 | AR | 25 | 311 | 0 | 1 |
| PR | Mangueirinha | Rheumatoid arthritis | 180300 | MF | 13 | 80 | 0 | 0 |
| SC | Criciúma | Growth hormone insensitivity with immunodeficiency | 245590 | AR | 61 | 759 | 0 | 0 |
| RS | Cândido Godói | Twinning | | MF | 0 | 50 | 0 | 0 |
| GO | Faina | Xeroderma pigmentosum | 278730 | AR | 0 | 15 | 0 | 0 |
| | **total** | | | | 687 | 12562 | 11 | 14 |

[a]MIM = Mendelian Inheritance in Man (https://www.ncbi.nlm.nih.gov/omim)

AD = autosomal dominant

AR = autosomal recessive

MF = multifactorial

NI = not identified

**Supplementary Table 9.** Medical geneticists (MGs) working at public healthcare system facilities.

| SUS facility | MGs 2019 (N = 208) | | MGs 2020 (N = 211) | |
|---|---|---|---|---|
| | N | % | N | % |
| health centre/basic health unit | 1 | 0.5 | 0 | 0.0 |
| specialized/ambulatory clinic | 25 | 12.0 | 19 | 9.0 |
| consultant's office | 16 | 7.7 | 17 | 8.1 |
| health cooperative | 2 | 1.0 | 3 | 1.4 |
| specialized hospital | 25 | 12.0 | 29 | 13.7 |
| general hospital | 104 | 50.0 | 110 | 52.1 |
| day hospital | 2 | 1.0 | 2 | 1.0 |
| polyclinic | 21 | 10.1 | 21 | 10.0 |
| diagnostic and therapeutic support service unit | 12 | 5.8 | 9 | 4.3 |
| mixed health unit | 0 | 0.0 | 1 | 0.5 |

**Supplementary Table 10.** Reference centres for rare diseases listed in the Many of Us are Rare website (accessed July 2021) and the Ministry of Health website (accessed October 2021).

| region | state | municipality | reference centres (N = 88) |
|---|---|---|---|
| North | Acre | Rio Branco | 3 |
| North | Pará | Belém | 2 |
| total North | | | 5 |
| Northeast | Maranhão | São Luís | 1 |
| Northeast | Ceará | Fortaleza | 4 |
| Northeast | Ceará | Juazeiro do Norte | 1 |
| Northeast | Rio Grande do Norte | Paranamirim | 1 |
| Northeast | Paraíba | Campina Grande | 1 |
| Northeast | Paraíba | João Pessoa | 1 |
| Northeast | Pernambuco | Recife | 2 |
| Northeast | Alagoas | Maceió | 2 |
| Northeast | Sergipe | Aracaju | 1 |
| Northeast | Bahia | Salvador | 3 |
| total Northeast | | | 17 |
| Southeast | Minas Gerais | Alfenas | 1 |
| Southeast | Minas Gerais | Barbacena | 1 |
| Southeast | Minas Gerais | Belo Horizonte | 3 |
| Southeast | Minas Gerais | Uberlândia | 1 |
| Southeast | Espirito Santo | Serra | 1 |
| Southeast | Espirito Santo | Vitória | 4 |

| | | | |
|---|---|---|---|
| Southeast | Rio de Janeiro | Duque de Caxias | 1 |
| Southeast | Rio de Janeiro | Niterói | 1 |
| Southeast | Rio de Janeiro | Petrópolis | 1 |
| Southeast | Rio de Janeiro | Rio de Janeiro | 15 |
| Southeast | Rio de Janeiro | Teresópolis | 1 |
| Southeast | Rio de Janeiro | Volta Redonda | 1 |
| Southeast | São Paulo | Bauru | 1 |
| Southeast | São Paulo | Campinas | 3 |
| Southeast | São Paulo | Osasco | 1 |
| Southeast | São Paulo | Ribeirão Preto | 1 |
| Southeast | São Paulo | Santo André | 1 |
| Southeast | São Paulo | Santos | 1 |
| Southeast | São Paulo | São Carlos | 1 |
| Southeast | São Paulo | São José do Rio Preto | 1 |
| Southeast | São Paulo | São Paulo | 8 |
| total Southeast | | | 49 |
| South | Paraná | Curitiba | 5 |
| South | Paraná | Londrina | 1 |
| South | Paraná | Maringá | 1 |
| South | Santa Catarina | Florianópolis | 2 |
| South | Rio Grande do Sul | Porto Alegre | 2 |
| South | Rio Grande do Sul | Rio Grande | 1 |
| South | Rio Grande do Sul | Santa Maria | 1 |
| total South | | | 13 |
| Centre-West | Distrito Federal | Brasília | 2 |
| Centre-West | Goias | Anápolis | 1 |
| Centre-West | Mato Grosso | Cuiabá | 1 |
| total Centre-West | | | 4 |

**Supplementary Table 11.** Municipalities with at least one medical geneticist reported by the medical demography study[1], one medical geneticist in a SUS facility, or a reference centre for rare diseases.

| municipality code | municipality |
|---|---|
| 110020 | Porto Velho |
| 120040 | Rio Branco |
| 130260 | Manaus |
| 150130 | Barcarena |
| 150140 | Belém |
| 160020 | Calçoene |
| 211130 | São Luís |
| 221100 | Teresina |
| 230370 | Caucaia |
| 230440 | Fortaleza |
| 230730 | Juazeiro do Norte |
| 240325 | Paranamirim |
| 240810 | Natal |
| 250400 | Campina Grande |
| 250750 | João Pessoa |
| 260290 | Cabo de Santo Agostinho |
| 260630 | Granito |
| 261160 | Recife |
| 270430 | Maceió |
| 280030 | Aracaju |
| 291080 | Feira de Santana |
| 292730 | Salinas da Margarida |
| 292740 | Salvador |
| 310160 | Alfenas |
| 310560 | Barbacena |
| 310610 | Belmiro Braga |
| 310620 | Belo Horizonte |
| 310670 | Betim |
| 312770 | Governador Valadares |
| 313240 | Itajubá |
| 313670 | Juiz de Fora |
| 313760 | Lagoa Santa |
| 313940 | Manhuaçu |
| 314480 | Nova Lima |
| 317010 | Uberaba |
| 317020 | Uberlândia |
| 320120 | Cachoeiro de Itapemirim |

| | |
|---|---|
| 320500 | Serra |
| 320520 | Vila Velha |
| 320530 | Vitória |
| 330060 | Bom Jesus do Itabapoana |
| 330100 | Campos dos Goytacazes |
| 330170 | Duque de Caxias |
| 330240 | Macaé |
| 330330 | Niterói |
| 330340 | Nova Friburgo |
| 330390 | Petrópolis |
| 330452 | Rio das Ostras |
| 330455 | Rio de Janeiro |
| 330580 | Teresópolis |
| 330630 | Volta Redonda |
| 350550 | Barretos |
| 350600 | Bauru |
| 350750 | Botucatu |
| 350945 | Campina do Monte Alegre |
| 350950 | Campinas |
| 351860 | Guariba |
| 352560 | João Ramalho |
| 352585 | Jumirim |
| 352590 | Jundiaí |
| 353430 | Orlândia |
| 353440 | Osasco |
| 353650 | Paulínia |
| 353870 | Piracicaba |
| 353900 | Pirangi |
| 354140 | Presidente Prudente |
| 354260 | Registro |
| 354340 | Ribeirão Preto |
| 354780 | Santo André |
| 354850 | Santos |
| 354870 | São Bernardo do Campo |
| 354880 | São Caetano do Sul |
| 354890 | São Carlos |
| 354940 | São Joaquim da Barra |
| 354980 | São José do Rio Preto |
| 354990 | São José dos Campos |
| 355020 | São Miguel Arcanjo |
| 355030 | São Paulo |
| 355170 | Sertãozinho |
| 355220 | Sorocaba |
| 355400 | Tatuí |
| 355410 | Taubaté |

| | |
|---|---|
| 355670 | Vinhedo |
| 410370 | Cambé |
| 410400 | Campina Grande do Sul |
| 410690 | Curitiba |
| 410830 | Foz do Iguaçu |
| 411370 | Londrina |
| 411520 | Maringá |
| 420240 | Blumenau |
| 420460 | Criciúma |
| 420535 | Flor do Sertão |
| 420540 | Florianópolis |
| 420900 | Joaçaba |
| 420910 | Joinville |
| 430160 | Bagé |
| 430460 | Canoas |
| 430760 | Estância Velha |
| 431140 | Lajeado |
| 431407 | Passo do Sobrado |
| 431410 | Passo Fundo |
| 431440 | Pelotas |
| 431480 | Portão |
| 431490 | Porto Alegre |
| 431560 | Rio Grande |
| 431690 | Santa Maria |
| 500270 | Campo Grande |
| 510340 | Cuiabá |
| 520110 | Anápolis |
| 520870 | Goiânia |
| 530010 | Brasília |

1. Scheffer M. *Demografía Médica No Brasil 2020.*; 2020.

**Supplementary Table 12.** Distribution of rare genetic diseases (RGD) live births and hospitalizations, and hereditary cancer syndrome (HCS) deaths in the public healthcare system, and medical geneticists (MGs), by municipality (2019-2020).

| region | state | municipality | RGD live births 2019-2020 (N = 53,746) | RGD hospital admissions 2019-2020 (N = 192,407) | HCS deaths 2019 (N = 229) | MGs[a] (N = 330) |
|---|---|---|---|---|---|---|
| North | Acre | Rio Branco | 144 | 1,050 | 2 | 1 |
| North | Amazonas | Manaus | 444 | 4,091 | 8 | 1 |
| North | Pará | Belém | 242 | 2,611 | 10 | 3 |
| Northeast | Maranhão | São Luís | 240 | 3,255 | 13 | 1 |
| Northeast | Piauí | Teresina | 227 | 2,575 | 5 | 1 |
| Northeast | Ceará | Caucaia | 135 | 831 | 1 | 2 |
| Northeast | Ceará | Fortaleza | 1,090 | 9,505 | 5 | 7 |
| Northeast | Rio Grande do Norte | Natal | 235 | 3,105 | 1 | 1 |
| Northeast | Paraíba | Campina Grande | 94 | 929 | 2 | 2 |
| Northeast | Paraíba | João Pessoa | 262 | 1,899 | 2 | 4 |
| Northeast | Pernambuco | Recife | 532 | 7,252 | 8 | 5 |
| Northeast | Alagoas | Maceió | 305 | 2,076 | 3 | 5 |
| Northeast | Sergipe | Aracaju | 206 | 1,115 | 4 | 5 |
| Northeast | Bahia | Salvador | 791 | 9,131 | 12 | 11 |
| Southeast | Minas Gerais | Belo Horizonte | 587 | 8,918 | 7 | 21 |
| Southeast | Minas Gerais | Betim | 115 | 1,185 | 2 | 1 |
| Southeast | Minas Gerais | Governador Valadares | 95 | 581 | 1 | 1 |
| Southeast | Minas Gerais | Juiz de Fora | 160 | 1,680 | 2 | 1 |

| | | | | | | |
|---|---|---|---|---|---|---|
| Southeast | Minas Gerais | Lagoa Santa | 25 | 211 | 1 | 1 |
| Southeast | Minas Gerais | Manhuaçu | 5 | 332 | 0 | 1 |
| Southeast | Minas Gerais | Nova Lima | 22 | 314 | 0 | 3 |
| Southeast | Minas Gerais | Uberaba | 58 | 1,040 | 3 | 1 |
| Southeast | Minas Gerais | Uberlândia | 147 | 5,142 | 2 | 2 |
| Southeast | Espirito Santo | Cachoeiro de Itapemirim | 51 | 887 | 0 | 1 |
| Southeast | Espirito Santo | Vila Velha | 149 | 1,165 | 3 | 3 |
| Southeast | Espirito Santo | Vitória | 135 | 1,099 | 2 | 2 |
| Southeast | Rio de Janeiro | Bom Jesus do Itabapoana | 6 | 233 | 0 | 1 |
| Southeast | Rio de Janeiro | Niterói | 94 | 1,188 | 4 | 2 |
| Southeast | Rio de Janeiro | Petrópolis | 117 | 814 | 0 | 1 |
| Southeast | Rio de Janeiro | Rio de Janeiro | 1,102 | 16,332 | 19 | 34 |
| Southeast | Rio de Janeiro | Volta Redonda | 44 | 528 | 1 | 1 |
| Southeast | São Paulo | Barretos | 15 | 483 | 1 | 1 |
| Southeast | São Paulo | Botucatu | 56 | 807 | 0 | 2 |
| Southeast | São Paulo | Campinas | 225 | 4,046 | 5 | 13 |
| Southeast | São Paulo | Guariba | 14 | 143 | 0 | 1 |
| Southeast | São Paulo | Jundiaí | 243 | 2,124 | 1 | 1 |
| Southeast | São Paulo | Paulínia | 39 | 352 | 0 | 2 |

| | | | | | | |
|---|---|---|---|---|---|---|
| Southeast | São Paulo | Piracicaba | 118 | 959 | 1 | 1 |
| Southeast | São Paulo | Pirangi | 2 | 46 | 0 | 1 |
| Southeast | São Paulo | Presidente Prudente | 62 | 959 | 1 | 1 |
| Southeast | São Paulo | Registro | 18 | 218 | 0 | 1 |
| Southeast | São Paulo | Ribeirão Preto | 296 | 2,934 | 2 | 13 |
| Southeast | São Paulo | São Bernardo do Campo | 236 | 2,303 | 1 | 1 |
| Southeast | São Paulo | São Caetano do Sul | 31 | 512 | 0 | 1 |
| Southeast | São Paulo | São Carlos | 10 | 1,823 | 1 | 1 |
| Southeast | São Paulo | São Joaquim da Barra | 7 | 143 | 0 | 1 |
| Southeast | São Paulo | São Paulo | 6,695 | 35,418 | 40 | 70 |
| Southeast | São Paulo | Sorocaba | 200 | 1,261 | 1 | 1 |
| Southeast | São Paulo | Vinhedo | 25 | 311 | 0 | 1 |
| South | Paraná | Cambé | 23 | 481 | 0 | 1 |
| South | Paraná | Curitiba | 255 | 8,359 | 8 | 13 |
| South | Paraná | Foz do Iguaçu | 72 | 1,050 | 2 | 1 |
| South | Paraná | Maringá | 84 | 1,304 | 2 | 1 |
| South | Santa Catarina | Blumenau | 67 | 1,208 | 1 | 1 |
| South | Santa Catarina | Florianópolis | 126 | 2,103 | 2 | 3 |
| South | Santa Catarina | Joinville | 131 | 2,449 | 1 | 2 |
| South | Rio Grande do Sul | Bagé | 10 | 349 | 0 | 1 |

| | | | | | | |
|---|---|---|---|---|---|---|
| South | Rio Grande do Sul | Canoas | 94 | 1,581 | 3 | 1 |
| South | Rio Grande do Sul | Estância Velha | 12 | 128 | 0 | 1 |
| South | Rio Grande do Sul | Lajeado | 42 | 185 | 0 | 1 |
| South | Rio Grande do Sul | Pelotas | 55 | 964 | 0 | 2 |
| South | Rio Grande do Sul | Porto Alegre | 399 | 6,690 | 1 | 33 |
| South | Rio Grande do Sul | Santa Maria | 71 | 771 | 2 | 1 |
| Centre-West | Mato Grosso do Sul | Campo Grande | 265 | 2,056 | 3 | 4 |
| Centre-West | Mato Grosso | Cuiabá | 130 | 1,180 | 3 | 1 |
| Centre-West | Goiás | Goiânia | 314 | 3,738 | 12 | 3 |
| Centre-West | Distrito Federal | Brasília | 732 | 11,895 | 12 | 21 |

[a]Two MGs did not provide municipality location.

**Supplementary Table 13.** Municipalities without a registered medical geneticist and a thousand or more hospital admissions for an RGD in 2019-2020.

| region | state | municipality | RGD hospital admissions 2019-2020 | HCS deaths 2019 | RGD live births 2019-2020 |
|---|---|---|---|---|---|
| Southeast | São Paulo | Guarulhos | 3,695 | 2 | 554 |
| Southeast | Minas Gerais | São Sebastião do Paraíso | 3,359 | 0 | 4 |
| Northeast | Pernambuco | Jaboatão dos Guararapes | 2,976 | 1 | 225 |
| Southeast | São Paulo | Santo André | 2,668 | 2 | 187 |
| Northeast | Ceará | Tauá | 2,611 | 0 | 8 |
| South | Paraná | Londrina | 2,325 | 4 | 152 |
| Southeast | Minas Gerais | Contagem | 2,307 | 2 | 89 |
| Southeast | Rio de Janeiro | Duque de Caxias | 2,196 | 3 | 268 |
| Northeast | Pernambuco | Olinda | 2,075 | 1 | 102 |
| North | Rondônia | Porto Velho | 2,064 | 1 | 166 |
| Southeast | São Paulo | São José do Rio Preto | 1,958 | 0 | 206 |
| Southeast | São Paulo | São José dos Campos | 1,894 | 2 | 205 |
| Southeast | Rio de Janeiro | São Gonçalo | 1,827 | 2 | 130 |
| Southeast | Rio de Janeiro | Nova Iguaçu | 1,731 | 1 | 117 |
| Northeast | Bahia | Feira de Santana | 1,707 | 3 | 55 |
| Southeast | São Paulo | Socorro | 1,658 | 0 | 4 |
| Southeast | São Paulo | Osasco | 1,643 | 3 | 217 |
| South | Paraná | São José dos Pinhais | 1,641 | 2 | 57 |
| Southeast | São Paulo | Louveira | 1,548 | 0 | 25 |
| Southeast | São Paulo | Barueri | 1,542 | 2 | 109 |
| South | Paraná | Ponta Grossa | 1,536 | 2 | 75 |
| North | Pará | Ananindeua | 1,492 | 5 | 56 |
| Northeast | Bahia | Vitória da Conquista | 1,427 | 2 | 76 |

| | | | | | |
|---|---|---|---|---|---|
| Southeast | São Paulo | Diadema | 1,421 | 1 | 313 |
| Northeast | Bahia | Ilhéus | 1,419 | 1 | 20 |
| North | Roraima | Boa Vista | 1,412 | 0 | 127 |
| Southeast | São Paulo | Carapicuíba | 1,407 | 2 | 114 |
| Southeast | São Paulo | Mogi das Cruzes | 1,378 | 1 | 114 |
| Southeast | Rio de Janeiro | São João de Meriti | 1,348 | 0 | 86 |
| Northeast | Pernambuco | Paulista | 1,322 | 1 | 57 |
| South | Paraná | Colombo | 1,278 | 0 | 63 |
| Southeast | Espirito Santo | Serra | 1,272 | 0 | 219 |
| Southeast | Rio de Janeiro | Belford Roxo | 1,238 | 1 | 100 |
| Southeast | Espirito Santo | Cariacica | 1,233 | 4 | 131 |
| Centre-West | Goiás | Aparecida de Goiânia | 1,179 | 1 | 113 |
| Southeast | Minas Gerais | Montes Claros | 1,160 | 1 | 108 |
| Southeast | Rio de Janeiro | Campos dos Goytacazes | 1,138 | 1 | 163 |
| Northeast | Bahia | Itabuna | 1,114 | 0 | 39 |
| Northeast | Pernambuco | Caruaru | 1,104 | 1 | 150 |
| South | Santa Catarina | Itajaí | 1,104 | 1 | 75 |
| Southeast | São Paulo | Embu das Artes | 1,101 | 1 | 77 |
| Southeast | São Paulo | Cotia | 1,090 | 0 | 81 |
| Southeast | Minas Gerais | Ipatinga | 1,083 | 1 | 60 |
| South | Santa Catarina | São José | 1,053 | 1 | 53 |
| South | Rio Grande do Sul | Caxias do Sul | 1,042 | 1 | 91 |
| Southeast | Minas Gerais | Ribeirão das Neves | 1,034 | 0 | 85 |
| Southeast | São Paulo | Franca | 1,022 | 2 | 112 |
| South | Paraná | Campo Largo | 1,020 | 2 | 23 |
| Southeast | Rio de Janeiro | Valença | 1,015 | 0 | 9 |
| Southeast | São Paulo | Bauru | 1,001 | 1 | 103 |

RGD = rare genetic diseases

HCS = hereditary cancer syndromes

**Supplementary Table 14.** Attention for rare genetic diseases (RGD) provided by public healthcare system facilities.

| **SUS facility** | **RGD 2019-2020** (N = 264,392) | **%** |
|---|---|---|
| centre for health and family support | 44 | 0,02 |
| centre for hemotherapy and/or haematological care | 829 | 0,31 |
| natural birth centre | 15 | 0,01 |
| health centre/basic health unit | 15,078 | 5,70 |
| state centre for notification, collection and distribution | 62 | 0,02 |
| specialized clinic/ambulatory | 19,221 | 7,27 |
| consultant's office | 562 | 0,21 |
| specialized hospital | 5,888 | 2,23 |
| general hospital | 49,058 | 18,56 |
| day hospital | 445 | 0,17 |
| central public health laboratory | 314 | 0,12 |
| public health laboratory | 5,707 | 2,16 |
| polyclinic | 8,351 | 3,16 |
| orthopaedic clinic | 38 | 0,01 |
| health centre | 532 | 0,20 |
| emergency care | 1,474 | 0,56 |
| specialized first-aid centre | 134 | 0,05 |
| general first-aid centre | 506 | 0,19 |
| home care | 56 | 0,02 |
| indigenous health care unit | 48 | 0,02 |
| diagnostic and therapeutic support service unit | 152,538 | 57,69 |
| mixed health unit | 3,410 | 1,29 |
| mobile river health unit | 48 | 0,02 |
| mobile land health unit | 34 | 0,01 |
